# Crowdfunding for health research: A global systematic review, qualitative evidence synthesis and TDR pilot for LMIC researchers

**DOI:** 10.1101/2021.11.08.21266070

**Authors:** Eneyi E. Kpokiri, Clarisse Sri-Pathmanathan, Priyanka Shrestha, Sana Navaid, Teerawat Wiwatpanit, Asha Wijegunawardana, Mahmud Ali Umar, Debra Jackson, Jackeline Alger, Meghan A. Bohren, Mia Hoole, Meredith Labarda, Noel Juban, Pascal Launois, Weiming Tang, Beatrice Halpaap, Joseph D. Tucker

**Affiliations:** Clinical Research Department, Faculty of Infectious and Tropical Diseases, London School of Hygiene and Tropical Medicine, London, United Kingdom; Department of Global Health, University of Washington, Seattle, USA; Institute of Global Health and Infectious Diseases, University of North Carolina at Chapel Hill, Chapel Hill, North Carolina, USA; National Center for Genetic Engineering and Biotechnology, National Science and Technology Development Agency, Pathum Thani, Thailand; Department of Bioprocess Technology, Faculty of Technology, Rajarata University of Sri Lanka; Department of Biology, Kano University of Science and Technology, Wudil, Kano State, Nigeria; Faculty of Epidemiology and Population Health, London School of Hygiene and Tropical Medicine, London, United Kingdom; Department of Clinical Laboratory, Hospital Escuela; Faculty of Medical Sciences, Universidad Nacional Autónoma de Honduras; and Instituto de Enfermedades Infecciosas Parasitología Antonio Vidal; Tegucigalpa, Honduras; Gender and Women’s Health Unit, Centre for Health Equity, Melbourne School of Population and Global Health, University of Melbourne, Carlton, VIC, Australia; Because Stories, UK; Department of Medicine, University of the Philippines, Manila-School of Health Sciences, Leyte, Philippines; University of North Carolina Project-China, Guangzhou, China; TDR, The Special Programme for Research and Training in Tropical Diseases co-sponsored by UNICEF, UNDP, the World Bank and WHO, hosted at WHO, Geneva, Switzerland

**Keywords:** Crowdfunding, LMIC, qualitative research, systematic review

## Abstract

**Background:** Many low-and middle-income country (LMIC) researchers have disadvantages when applying for research grants. Crowdfunding may help LMIC researchers to fund their research. Crowdfunding organizes large groups of people to make small contributions to support a research study. This manuscript synthesizes global qualitative evidence and describes a TDR crowdfunding pilot for LMIC-based researchers.

**Methods:** Our global systematic review and qualitative evidence synthesis searched six databases for qualitative data. We used a thematic synthesis approach and assessed our findings using the GRADE-CERQual approach. Building on the review findings, we organized a crowdfunding pilot to support LMIC researchers and use crowdfunding. The pilot provided an opportunity to assess the feasibility of crowdfunding for infectious diseases of poverty research in resource-constrained settings.

**Results:** Nine studies were included in the qualitative evidence synthesis and we identified seven themes. Seven studies demonstrated that strong public engagement facilitated crowdfunding for research. Other themes included the correlates of crowdfunding success, risks of crowdfunding, and risk mitigation strategies. Our pilot data suggest that crowdfunding is feasible in diverse LMIC settings. Three researchers launched crowdfunding campaigns, met their goals and received substantial monetary (raising a total of $26,546 across all five campaigns) and non-monetary contributions. Two researchers are still preparing for campaign launch due to COVID-19 related difficulties.

**Conclusion:** Public engagement provides a foundation for effective crowdfunding for health research. Our evidence synthesis and pilot data provide practical strategies for LMIC researchers to engage the public and use crowdfunding. A practical guide was created alongside to facilitate these activities across multiple settings.

**What is already known?:** Crowdfunding has been used to fund health causes, technology start-ups, creative projects, and more recently, scientific research. Although crowdfunding has been used for research funding in high-income settings, there is less evidence from LMIC settings. In addition, previous reviews of crowdfunding have not focused on public engagement strategies that may be important for developing effective crowdfunding campaigns.

**What are the new findings?:**

- Our qualitative evidence synthesis finds that crowdfunding research focuses on creators and backers in high-income settings, neglecting LMIC researchers.
- The TDR pilot programme suggests that crowdfunding is feasible for LMIC researchers. Three of the five pilot finalists exceeded their crowdfunding goals and received substantial non-monetary contributions.

## Introduction

Crowdfunding engages large groups of people who make small contributions to support a research study.^1^ It provides a method for researchers to engage with the public to spur interest and cultivate local partnerships.^2^ From a public perspective, crowdfunding provides a way for people to invest in locally relevant topics and directly contribute to science. Crowdfunding has been used to support research studies in many high-income countries,^3-5^ but has not been used in low- and middle-income countries (LMIC).^6^

LMIC-based researchers are often disadvantaged in applying for research grants compared to their high-income country (HIC) counterparts due to power asymmetries within global health.^7^ A telling example of this is the imbalance in authorship and high-impact global health journals across the world.^8,9^ Another example is the ‘brain drain’ of LMIC expertise, where valuable, non-remunerated time is spent on grant applications to UK-based funding, with high rejection rates and a lack of feedback.^10^ Additionally, a lot of the research in LMICs is externally funded, potentially leading to donor-driven research agendas, with a disregard for local needs, knowledge, and languages.^11,12^ One potential way to expand LMIC research funding is crowdfunding, where researchers communicate and engage with their communities, raise money at the local and international level, in order to conduct meaningful research. More local funding for research is one step to disrupting the unequal relationships observed within global health and may contribute to creating ‘Southern’ networks thereby increasing LMIC ownership.^11^ In addition, local researchers working in their own communities may have a greater likelihood of transitioning to sustainable, locally financed public programs compared to foreign-funded research. Crowdfunding is an opportunity to democratise, decentralise and decolonise health research, and to build health networks between like-minded researchers and their communities.

We conducted a systematic review and qualitative evidence synthesis to identify practical suggestions for research crowdfunding. Because we could only find global or HIC-based evidence on crowdfunding, we organised a TDR Global open call and pilot programme to support selected LMIC researchers with their own crowdfunding for research campaigns. TDR, the Special Programme for Research and Training in Tropical Diseases is based at the World Health Organization (WHO) and is co-sponsored by the United Nations Children’s Fund (UNICEF), the United Nations Development Programme (UNDP), the World Bank and WHO. TDR Global is a community of passionate scientists and experts working to support global research efforts on infectious diseases of poverty. This manuscript synthesizes global qualitative evidence and describes a TDR pilot focused on public engagement and crowdfunding led by LMIC health researchers. The overall aim is to expand the literature by summarising the available evidence on crowdfunding for research and by assessing its feasibility in LMIC settings.

## Methods

### Systematic review and qualitative evidence synthesis

The purpose of this review was to synthesize evidence on crowdfunding for health research, including barriers, facilitators and implications for policy and practice. We initially used a scoping review approach, however we found rich qualitative data on crowdfunding for scientific research and the correlates of crowdfunding success, which prompted us to upgrade our methodology to a systematic review.^13^ We followed the Cochrane handbook for conducting systematic reviews and used 2020 PRISMA guidance.^14,15^ This systematic review was registered on the Open Science Framework platform (10.17605/OSF.IO/E7QH5).^16^

We searched PubMed, EMBASE, Web of Science, Scopus, Global Health and Google Scholar. We used the key terms [Crowdfunding or public funded or public contribution] OR AND [Research]. We also searched registers for grey literature including theses and dissertations, article pre-prints, conference proceedings, and the reference lists of relevant manuscripts. Search outputs from the databases were combined and deduplicated.

Search outputs were screened by title, then abstract and finally full text. Our inclusion criteria were limited to studies reporting crowdfunding in health research and published in English, between 1^st^ January 2000 and 23^rd^ March 2021. The search was updated on 22^nd^ September 2021. We included primary qualitative and mixed methods studies (where qualitative data were able to be extracted), whilst purely quantitative studies, editorials, opinion pieces, practical guides and reviews without qualitative data were excluded. Two independent reviewers (EK and CS) screened studies for inclusion and disagreements were resolved through consensus-based discussion with the wider team. EK and CS extracted relevant data, including study objectives, participants, study setting, study design, data collection methods, qualitative themes, main study findings and where possible, correlates of crowdfunding success.

We also independently assessed methodological limitations using the Critical Appraisal Skills Programme (CASP) tool with a checklist for each study, including validity, relevance, adequacy, methodological limitations and risk of bias.^17^

We used a thematic synthesis approach^18^ for data analysis, which involved familiarization with the data, coding the primary studies, developing themes, and using these themes to generate further understanding and hypotheses. We used the GRADE-CERQual (Confidence in the Evidence from Reviews of Qualitative research) approach to assess confidence in each qualitative review finding, based on four key components: methodological limitations, coherence of the review finding, adequacy of the data, and relevance of the included studies to the review question.^19-24^ After assessing each of these components, we made an overall judgement on the confidence we had in each review finding (high, moderate, low or very low). The CERQual approach has been applied to qualitative and mixed methods systematic reviews in a number of WHO global guidelines, because it provides high levels of transparency and precision.^19^

### The pilot programme

Building on the themes identified in the qualitative evidence synthesis, we developed a crowdfunding pilot programme in partnership with TDR Global. The main objective of this pilot programme was to test the effectiveness of crowdfunding as a means to finance health research in LMICs. The pilot took place in three stages: an open call to solicit LMIC researchers interested in crowdfunding; a capacity building workshop; launch of a crowdfunding campaign with mentorship and support for finalists (Figure 1).

**Figure 1:**
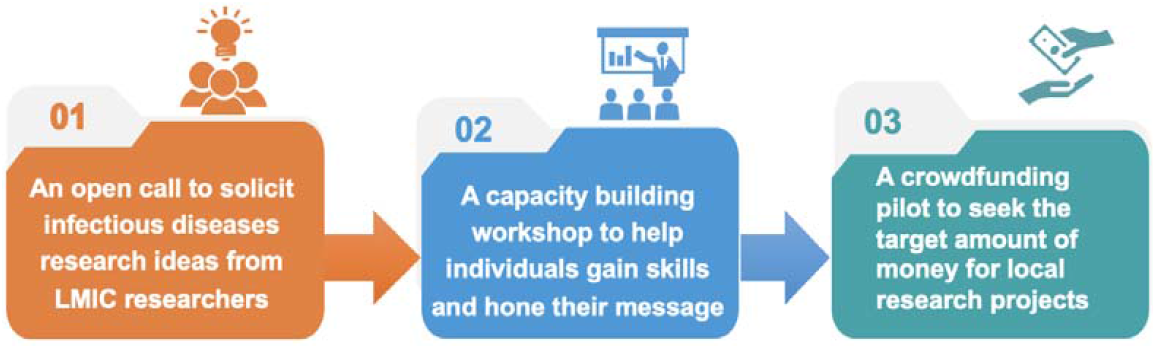
Stages of the TDR Global Pilot Programme.

### The open call

The crowdsourcing open call was designed using the framework provided by the TDR/SESH/SIHI Practical Guide on Crowdsourcing in Health and Health Research.^25^

Our open call was conducted in five steps, including convening a steering committee, promoting for community participation and engagement, receiving and judging contributions, recognising the finalists and implementing solutions. Detailed information on the process of these stages is described in Appendix I. We invited stakeholders in crowdfunding to join the steering committee. The crowdfunding call had 15 confirmed steering committee members (nine women and six men). All members of the steering committee had LMIC experience in crowdfunding for research or public engagement. The steering committee met monthly via a one-hour videoconference to provide guidance on the open call. The open call accepted submissions over six weeks. We promoted the call for submissions using infographics shared on social media channels and emails. At the end of the call, all submissions were screened for eligibility by the research team and eligible entries were sent to judges. We invited independent judges from the WHO TDR Global network to review submissions. Criteria for evaluation included compelling science, capacity for public engagement, and personal connection to the infectious disease topic. A total of 592 people volunteered to serve as judges and 47 were selected to review submissions. We selected volunteer judges based on TDR Global membership and LMIC research experience.

### Capacity building

The finalists were recognised through a TDR announcement and supported to attend a capacity building workshop in Geneva. The one and half day workshop included 1:1 mentoring from TDR Global members, presentations on crowdfunding (Appendix II), and group discussions about how to enhance public engagement and crowdfunding in LMICs. After the workshop, a monthly working group composed of finalists and mentors reported on crowdfunding progress.

### Campaign launch

Three finalists launched crowdfunding campaigns. They used multiple public engagement strategies to solicit both monetary and non-monetary contributions for their research projects. At the end of their campaigns, all three finalists exceeded their target amounts and raised between 7,000 and 11,000 USD.

A working group and end-user group, with professional and practical experience with crowdfunding for health research, were invited to comment on several drafts of this systematic review. In the final stages, a TDR Global external peer review was completed and six LMIC-based peer reviewers also provided feedback. A practical guide was developed by the same authors alongside this systematic review, using an adapted Delphi method to enable co-creation. It is available online and provides practical advice on how to organise public engagement and crowdfunding for health research, using evidence from this review. ^1^

## Results

### Qualitative evidence synthesis

This qualitative evidence synthesis summarizes evidence from published literature on facilitators and barriers of crowdfunding for research. Our initial electronic searches yielded 498 citations after deduplication (Figure 2). We assessed articles through title screening, abstract and finally through full-text screening. After exclusions, six papers from the database search met our inclusion criteria. An additional three studies were retrieved from reference lists and a grey literature search. Of the nine included studies, seven focused on high-income countries and two included global data, including from LMICs. Seven studies reported on data from one country and two reported on data from multiple countries. Four studies were qualitative studies and five were mixed methods. Full detail of the critical appraisal checklists completed for each study is available in Appendix VI.

**Figure 2:**
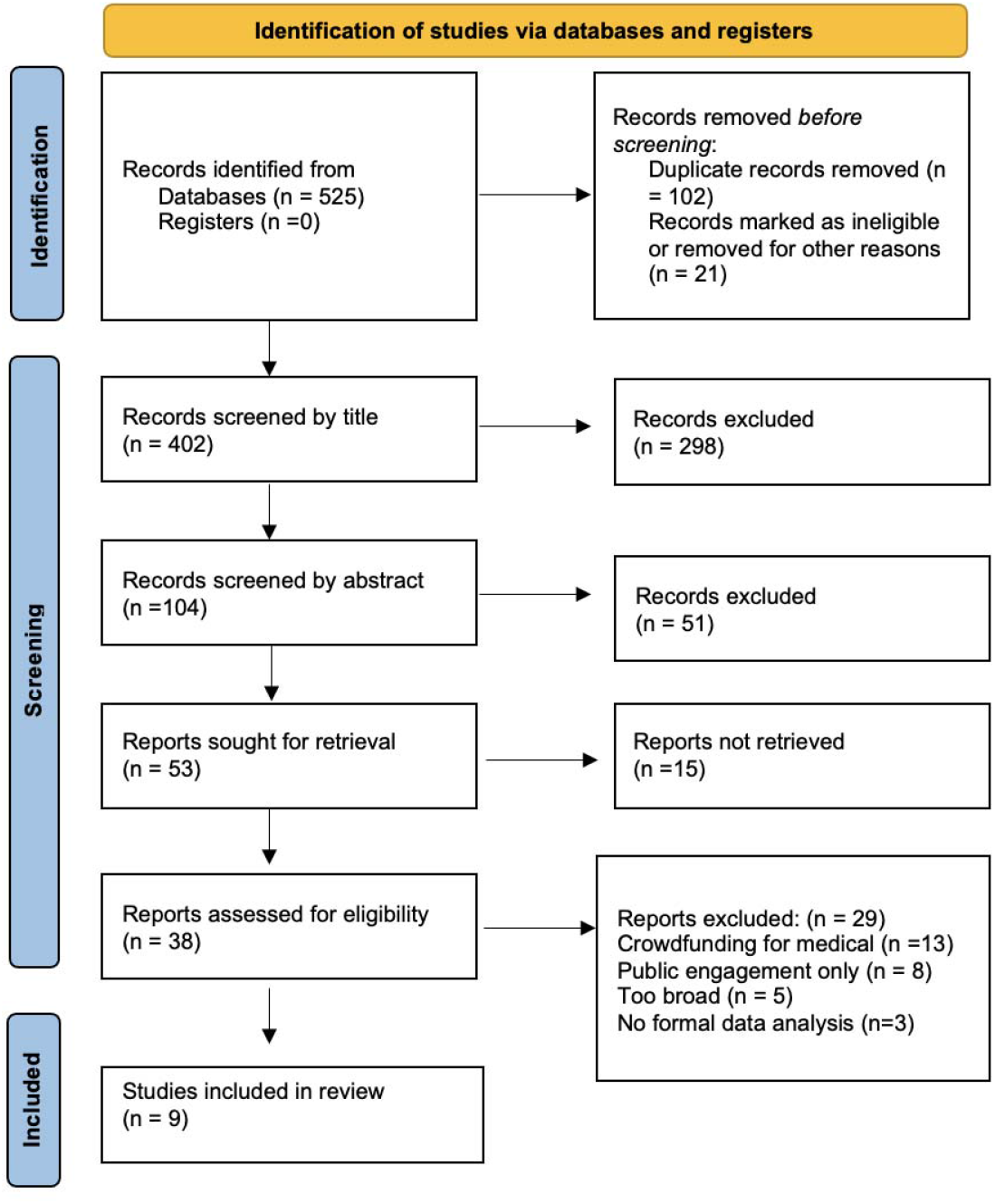
PRISMA flow diagram showing selection of studies^20^.

Of the nine included papers, six explored the processes and factors that were associated with successful crowdfunding campaigns.^26-29 2,30^ Three articles assessed the feasibility of conducting crowdfunding for health and/or scientific research.^31-33^ The characteristics of included studies and their main findings are provided in Appendix III. We identified seven findings which we organised into three broad domains: public engagement strategies, correlates of crowdfunding success, and risks and mitigation strategies. Of the seven findings, five were graded as moderate confidence and two were graded as low confidence using the GRADE-CERQual approach (Table 2).

**Table 1:**
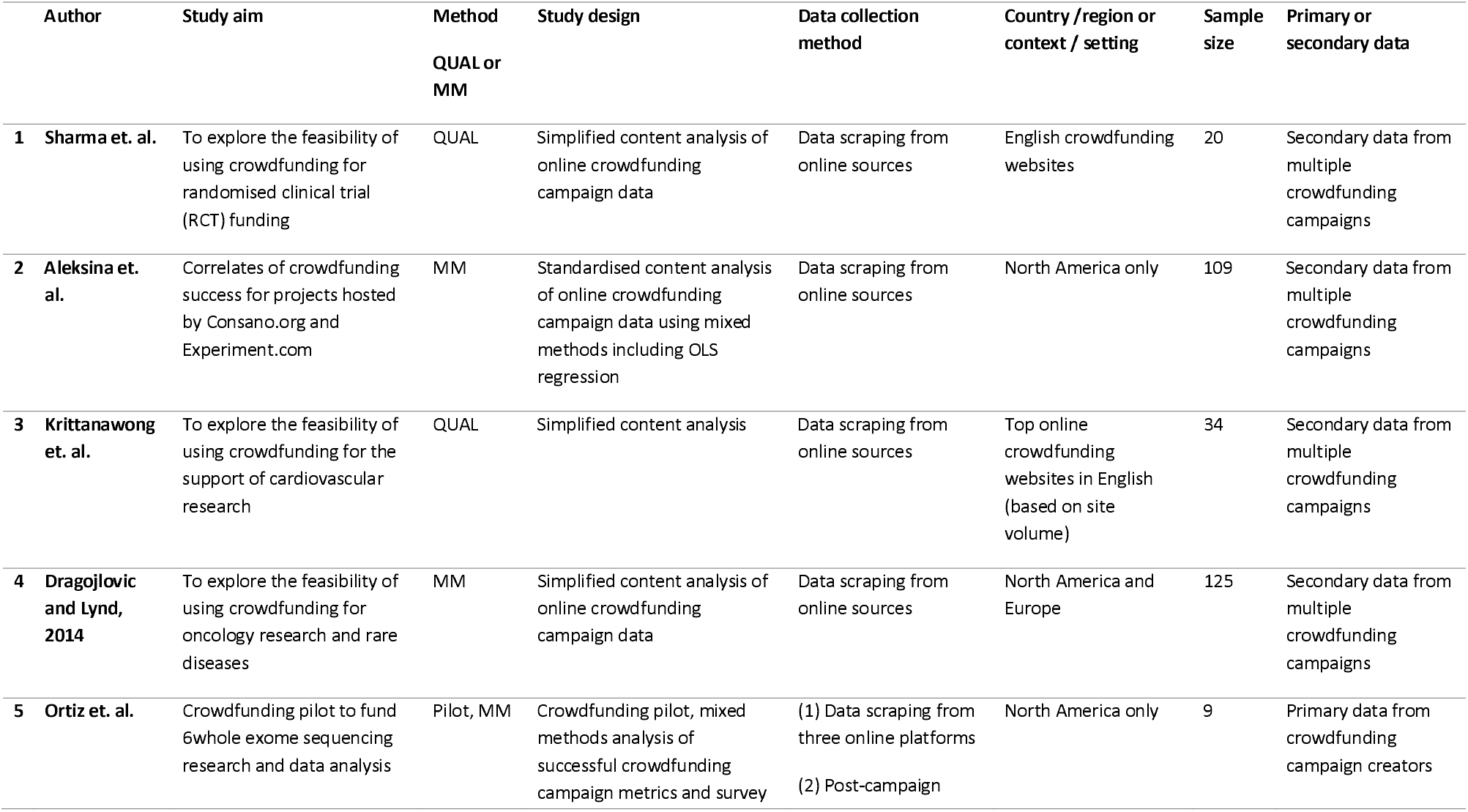
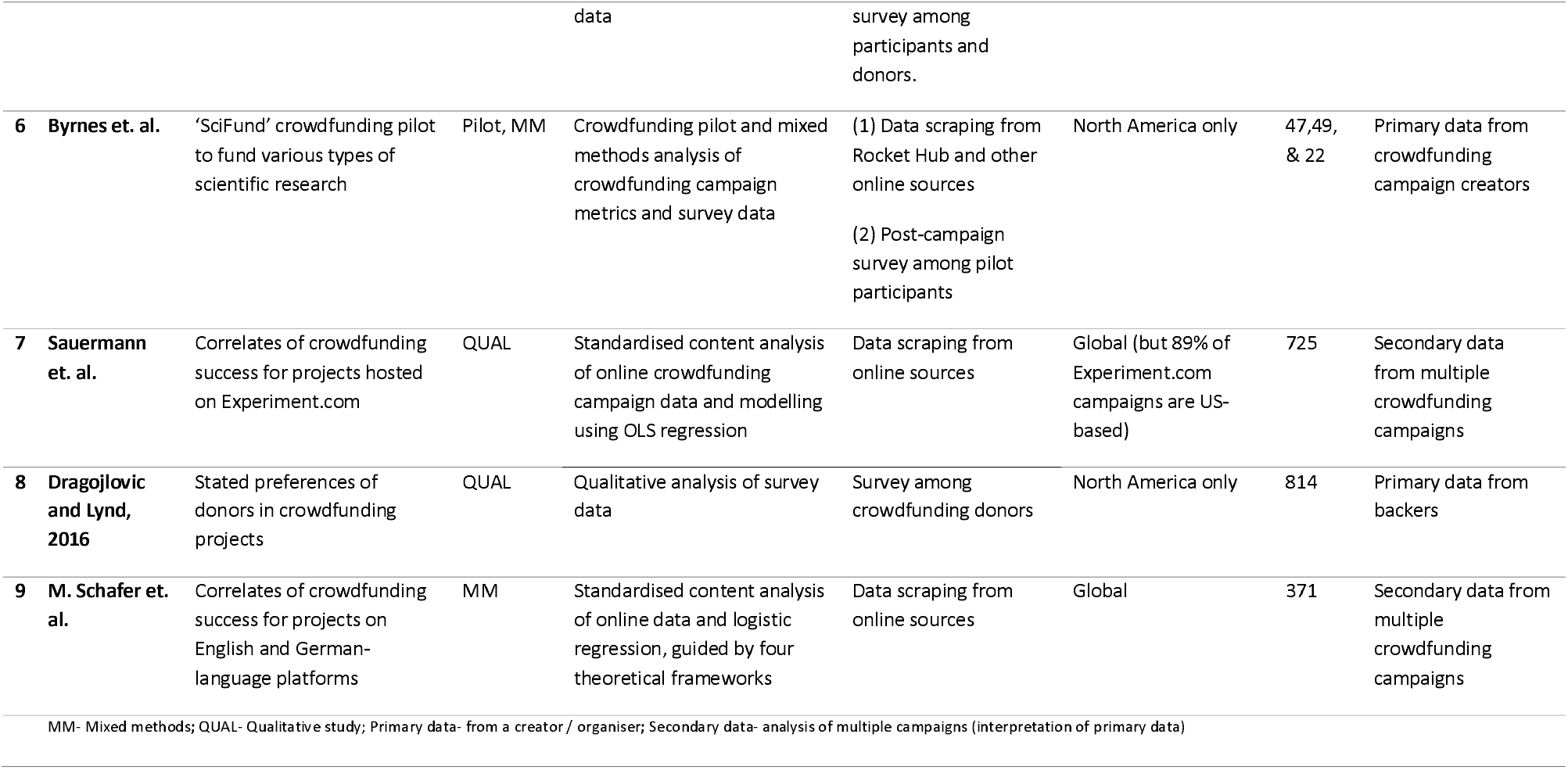
Summary of included studies and quality assessment.

**Table 2:**
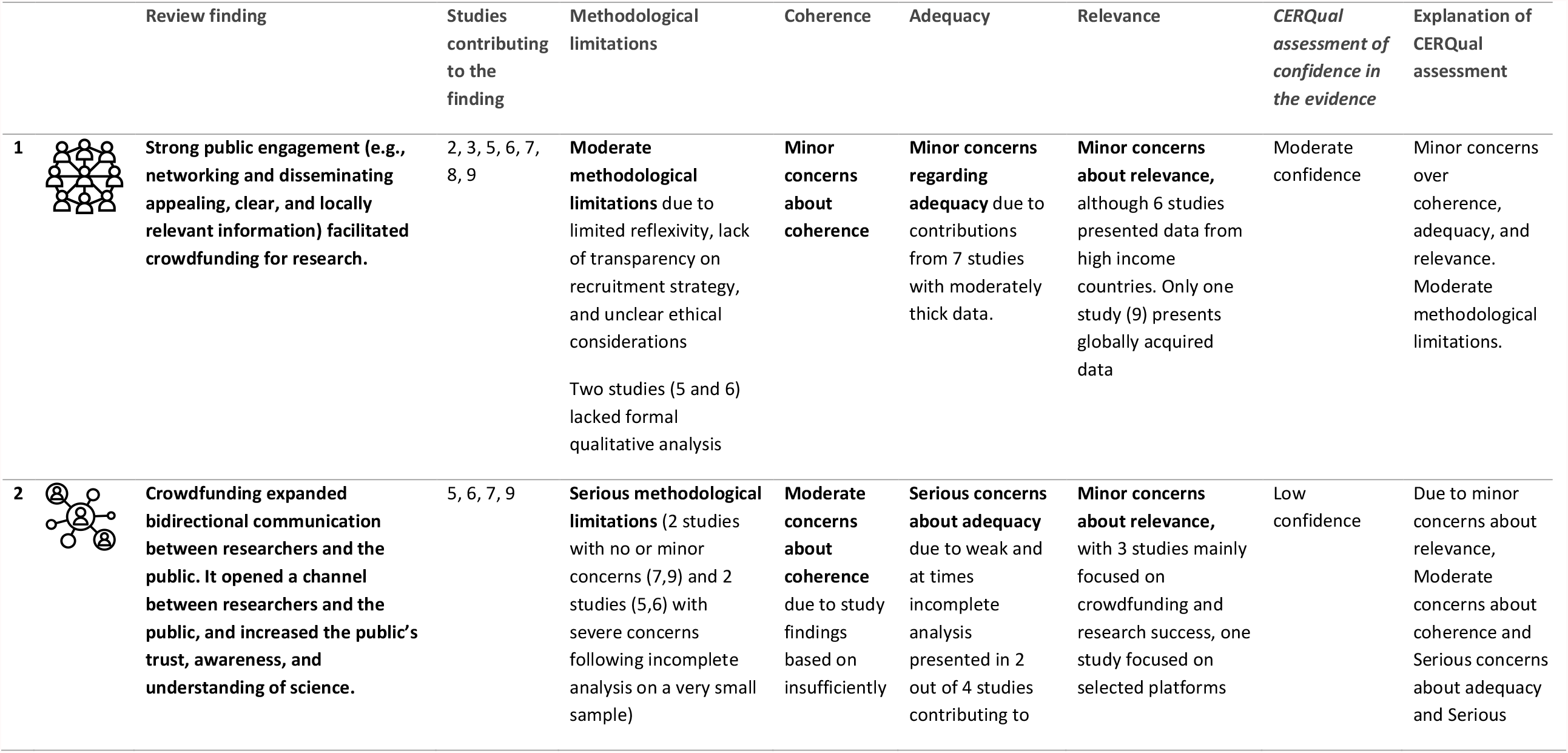
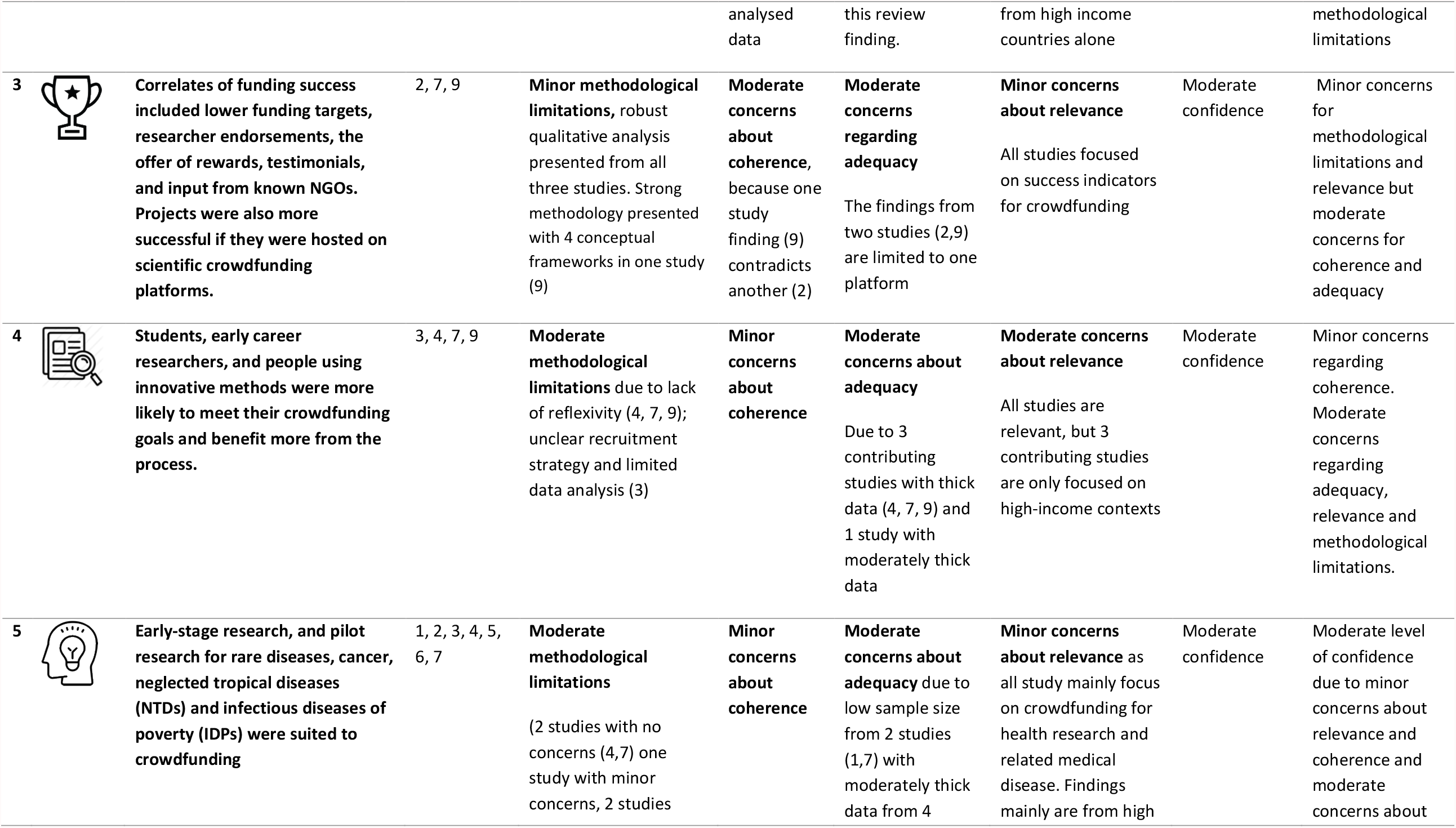
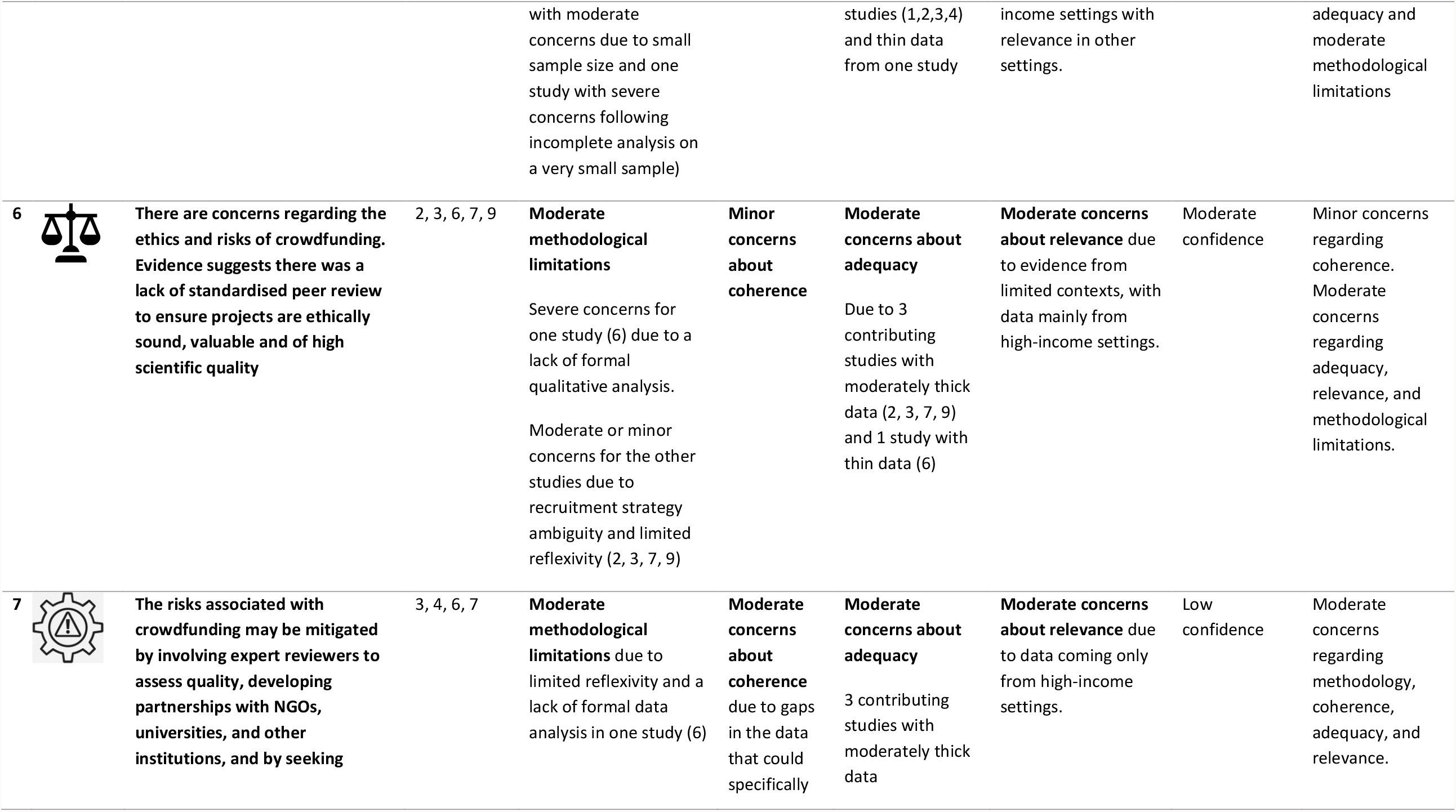
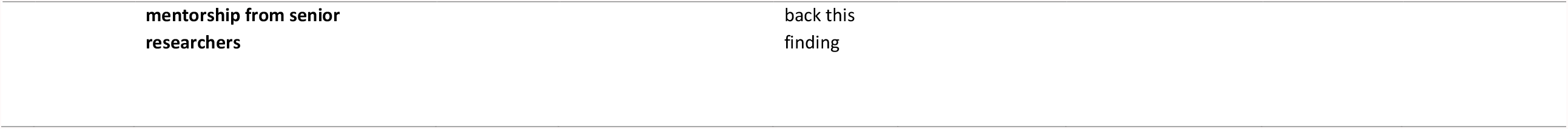
Evidence profile.

### Public engagement strategies

#### 1. Strong public engagement (e.g., networking and disseminating appealing, clear, and locally relevant information) facilitated crowdfunding for research. (Moderate confidence)^2,26-30,33^

We define public engagement in research as a two-way communication between the researcher and the public for mutual benefit. Evidence showed that networking skills and the ability to share a campaign among personal, professional, and social media networks were strongly associated with achieving crowdfunding campaign goals. Using multiple communication channels, including social media, blogs, direct contact, e-mail, newspaper, community radio, in-person events and conferences was also recommended. Using simple messages delivered by image or video increased donations : four studies suggested that campaigns with videos were more likely to succeed and were preferred by potential backers.^28-30,33^ They also found that keeping the audience updated through regular communication during and after the campaign led to more pledges and higher odds of success.^28,29^ Four studies found that researchers who partnered with non-governmental organizations, universities, or foundations enhanced their public engagement achievements.^26-28,31^

#### 2. Crowdfunding expanded bidirectional communication between researchers and the public. This channel between researchers and the public increased the public’s trust, awareness, and understanding of science. (Low confidence)^2,28-30^

One study found that feedback mechanisms, particularly two-way feedback between the backers and the researcher, significantly increased crowdfunding success.^30^ Evidence showed that crowdfunding can also help to bridge the gap between society and science by promoting public understanding of science through accessible resources. ^2,28-30^

### Correlates of crowdfunding success

#### 3. Correlates of funding success included lower funding targets, researcher endorsements, offer of rewards, testimonials, and input from known NGOs. Projects were also more successful if they were hosted on scientific crowdfunding platforms. (Moderate confidence)^27,29,30^

In addition to public engagement and communication strategies, certain factors were associated with crowdfunding success. One study found that campaigns hosted on specialized scientific crowdfunding platforms were more likely to reach their goals compared to campaigns on general interest crowdfunding platforms.^27^ Projects that offered rewards (e.g., small gifts to backers) had higher odds of achieving their goals.^29^ The evidence on researcher endorsements is mixed. One study found that researcher endorsements by other professionals increased funding success,^29^ but another found that research quality signals (highest academic title, scientific awards and the complexity and length of project description) had no effect on funding success.^30^ Similarly, endorsements and the sponsorship of platforms by established journals were not correlated with funding success. In a survey of stated preferences, one study found that researcher reputation is important to backers.^26^

#### 4. Students, early career researchers and people using innovative methods were more likely to meet their crowdfunding goals and benefit from the process. (Moderate confidence)^29-31,33^

Four studies found that students, early career researchers, and people with innovative studies were more likely to meet their campaign goals and benefit from the process.^29-31,33^ Early career researchers were defined as people within ten years of a terminal degree and it was found they had higher rates of achieving financial crowdfunding goals. Although established researchers have larger research networks, crowdfunding engages broader audiences, therefore traditional markers of quality, such as prior publications and researcher reputation, may not be so important. Three studies found that project risk was not associated with lower odds of success.^2,29,30^ However, one study found that some donors remained risk averse and that innovative projects were modestly less successful.^27^

#### 5. Early-stage research, and pilot research for rare diseases, cancer, neglected tropical diseases (NTDs) and infectious diseases of poverty (IDPs) were suited to crowdfunding. (Moderate confidence)2,27-29,31-33

Seven studies showed that crowdfunding may be an effective option to rapidly raise funds for research projects.^2,27-29,31-33^ Studies suggested that crowdfunding may be especially useful for pilot, phase 1 clinical trials or early-stage proof-of-concept research because campaigns with smaller targets were usually more successful. ^26,32^ Crowdfunding could complement or extend an existing research project. Alternatively, crowdfunding could support pilot studies before application to larger funding grants.^33^ One study on crowdfunding for clinical trials found that 95% of campaigns used a flexible model where researchers kept all the funds raised.^32^ These flexible models enabled researchers to get started on projects regardless of whether they reached their target, in contrast to all-or-nothing models, making crowdfunding a useful source of ‘seed money’. Two studies found crowdfunding is an effective way to support drug development on cancer, rare diseases, NTDs and IDPs.^28,31^

### Risks and mitigation strategies

#### 6. There were concerns regarding the ethics and potential risks of crowdfunding. Evidence suggested there was a lack of standardised peer review to ensure projects are ethically sound, valuable and of high scientific quality. (Moderate confidence)^2,27,29,30,33^

Five studies found that crowdfunding for scientific research was based on the public’s judgement and may thus promote research that is low-value, ethically unsound or not methodologically rigorous.^2,27,29,30,33^ Additional limitations of crowdfunding include the inability to monitor research funding allocation post-campaign and to sanction fraud and falsification.

#### 7. The risks associated with crowdfunding may be mitigated by involving expert reviewers to assess quality, developing partnerships with NGOs, universities, and other institutions, and seeking mentorship from senior researchers. (Low confidence)^2,29,31,33^

Two studies found an internal peer review system could be a solution to promoting high quality research related to crowdfunding.^29,33^ Some platforms required approval from ethical committees prior to launching their campaign, but these requirements varied.^29^ Seeking mentorship and partnering with NGOs specialised in marketing and fundraising helped researchers.^31^ They could facilitate efficient research administration and facilitate payment collection.

### Pilot programme

The open call received 121 unique submissions from researchers based in 37 LMICs. The judging process was conducted in three phases. In the first phase, all 121 entries were screened for eligibility using pre-defined criteria, including clear description of the scientific question and hypothesis, significance of the project and relevance to the public. This initial screening yielded 66 eligible entries. All eligible entries were then reviewed by judges and assigned scores. The five entries with a score of 7 and above were selected as finalists to receive detailed feedback and crowdfunding campaign support. The five finalists were from Guatemala, Mozambique, Nigeria, Sri Lanka, and Thailand (Table 3). All described social innovations in health and focused on infectious diseases of poverty.

**Table 3:**
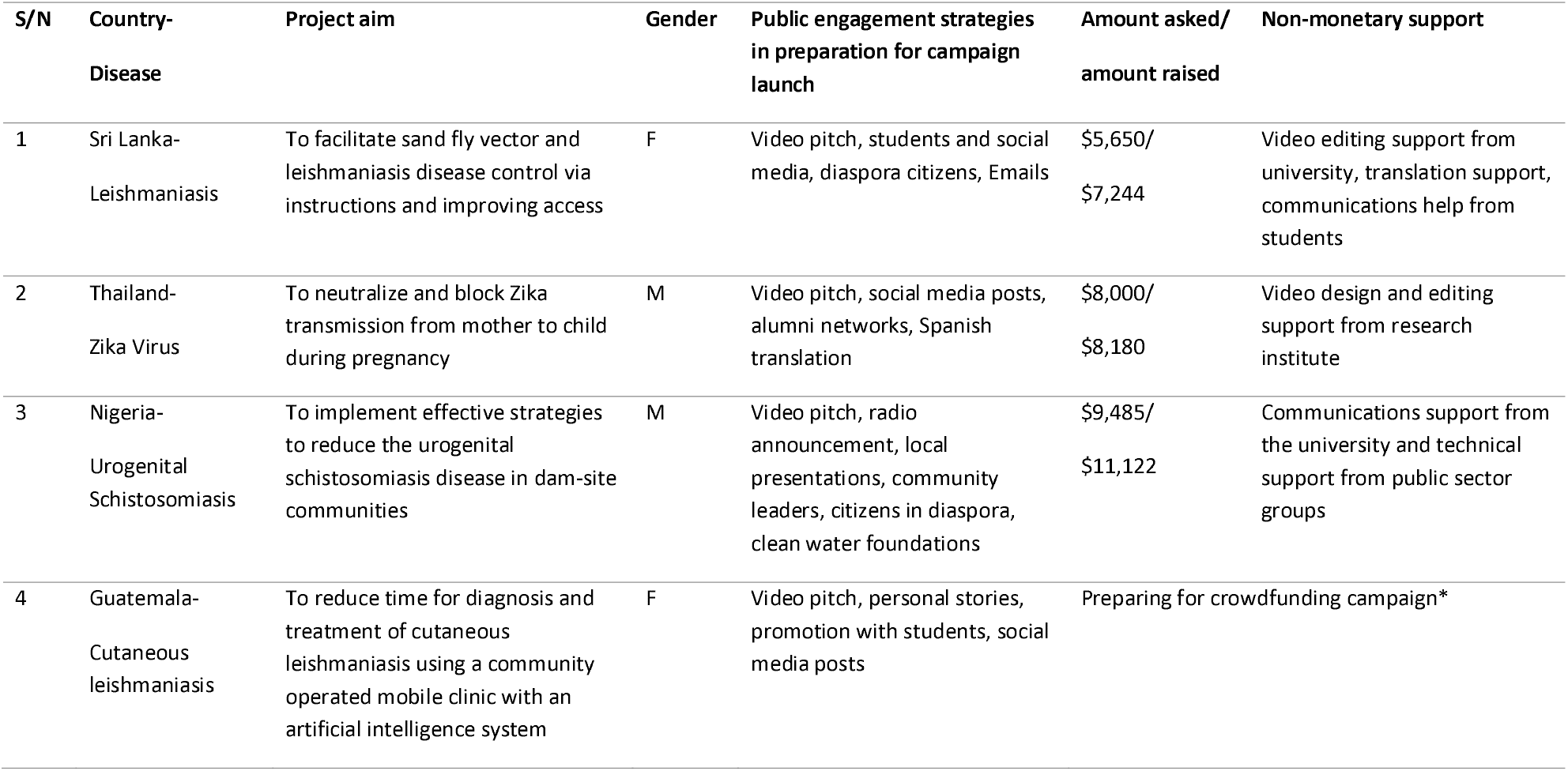
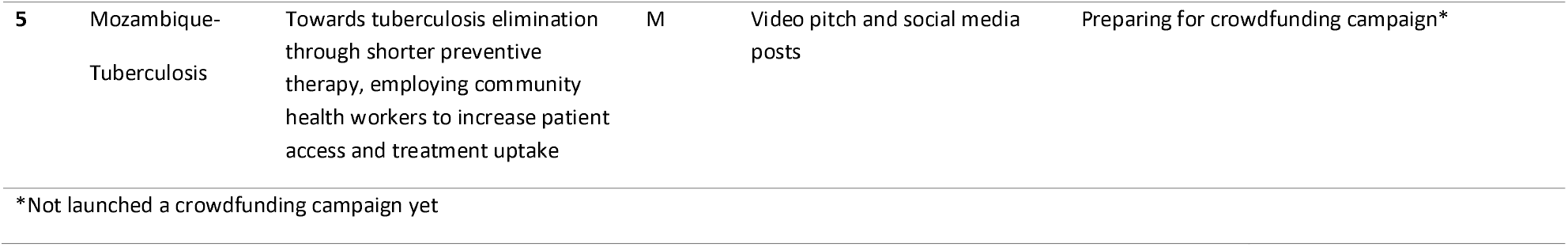
Details of finalist projects for public engagement and crowdfunding.

| **S/N** | **Country-**<br>**Disease** | **Project aim** | **Gender** | **Public engagement strategies**<br>**in preparation for campaign**<br>**launch** | **Amount asked/**<br>**amount raised** | **Non-monetary support** |
| --- | --- | --- | --- | --- | --- | --- |
| 1 | Sri Lanka-Leishmaniasis | To facilitate sand fly vector and leishmaniasis disease control via instructions and improving access | F | Video pitch, students and social media, diaspora citizens, Emails | \$5,650/<br>\$7,244 | Video editing support from university, translation support, communications help from students |
| 2 | Thailand-Zika Virus | To neutralize and block Zika transmission from mother to child during pregnancy | M | Video pitch, social media posts, alumni networks, Spanish translation | \$8,000/<br>\$8,180 | Video design and editing support from research institute |
| 3 | Nigeria-Urogenital<br>Schistosomiasis | To implement effective strategies to reduce the urogenital schistosomiasis disease in dam-site communities | M | Video pitch, radio announcement, local presentations, community leaders, citizens in diaspora, clean water foundations | \$9,485/<br>\$11,122 | Communications support from the university and technical support from public sector groups |
| 4 | Guatemala-Cutaneous<br>leishmaniasis | To reduce time for diagnosis and treatment of cutaneous leishmaniasis using a community operated mobile clinic with an artificial intelligence system | F | Video pitch, personal stories, promotion with students, social media posts | Preparing for crowdfunding campaign* |  |

|  |  |  |  |  |  |
| --- | --- | --- | --- | --- | --- |
| 5 | Mozambique-Tuberculosis | Towards tuberculosis elimination through shorter preventive therapy, employing community health workers to increase patient access and treatment uptake | M | Video pitch and social media posts | Preparing for crowdfunding campaign* |

\*Not launched a crowdfunding campaign yet

All five finalists used the tools of public engagement to develop campaign videos for their research project and benefitted from substantial non-monetary support (Table 3). In-kind contributions included assistance with developing and editing short videos from their universities, student support on social media, and scientific mentorship from TDR Global members. Among the five finalists, three launched crowdfunding campaigns. All three exceeded their original financial crowdfunding goals (Table 3).

The pilot program also identified potential risks and risk mitigation strategies (Appendix V). Potential risks of crowdfunding included fraud and deception, misinformation, unfair allocation of funds and lack of public interest in the project. Strategies to mitigate these risks included obtaining ethical approvals and support from local experts, clear communication throughout the campaign, sharing project results using open access tools, transparent engagement through videos and personal stories, and partnerships with universities or community-based organizations.

## Discussion

This paper expands the literature by summarising the available evidence on crowdfunding for health research and by assessing its feasibility in LMIC settings. Most of the evidence collected in our review has come from high-income country settings.^32,34-36^ The pilot programme complements this by demonstrating that LMIC researchers can benefit from the monetary and non-monetary support that crowdfunding provides. Crowdfunding could also be a powerful tool to decentralise and democratise research funding in resource-constrained settings.

Both the systematic review and pilot programme highlight that public engagement is essential for crowdfunding. Previous studies have shown that public engagement generates interest, which in turn leads to backers offering to help with projects and providing feedback.^37^ Public engagement skills may help to translate scientific concepts into more easily understood messages.^37^ Active engagement with the public during the campaign across a wide range of mediums (lab notes, e-mail updates, online webinars) can increase fundraising success.^29^ Although all three pilot programme finalists who launched their campaign had limited social media experience, they were successful in developing effective digital engagement strategies. These three finalists used videos as part of their campaigns - this may have enhanced the public’s trust in their projects, thus contributing to their crowdfunding success, consistent with evidence on the importance of videos in science communication.^38^ The finalists received training on storytelling, and they found that using personal stories from affected community members made their video pitches more meaningful. This is consistent with fundraising literature demonstrating that personal stories can be a useful tool to seek funding from donors for non-profit causes.^39,40^

Our systematic review shows that early-stage investigators and research studies with innovative methods were likely to reach crowdfunding goals. The public may place less emphasis on previous research experience compared to other research grant funding application processes.^27,30^ Therefore, campaigns with a broad engaged audience and efficient public engagement strategies alone can be successful in funding innovative research. In addition, we found that crowdfunding is useful for early-stage research and can then be used as preliminary data for larger grants.

Our pilot programme data demonstrates that crowdfunding is feasible in diverse LMICs settings. Evidence suggests there are barriers to seeking traditional research funding for many LMIC researchers, including fewer institutional research resources, less experience with research grants, and racism in science.^41,42^ One previously mentioned example is authorship and the fact that LMIC researchers who have worked in international partnerships are less likely to be first or corresponding authors.^43^ This likely disadvantages LMIC researchers when applying for grants as authorship in publications is often a marker of researcher reputation and signals productivity.^44^ Crowdfunding may be a useful tool for LMIC researchers to directly obtain support for research with less reliance on external donors or HIC researchers. It can also be argued that because crowdfunded research is often more grassroots and community-based, it may be more ethical and have a more enduring positive impact.^45^

Our data identified strategies to mitigate ethical issues associated with crowdfunding. We found mentorship from local experts could alleviate some of the concerns raised about the limited peer review of crowdfunded health research projects. During the pilot, our TDR Global team were involved in building local partnerships and mentorship opportunities to mitigate this risk. Additional risk mitigation strategies include obtaining ethical committee review approval prior to launch, ensuring transparency throughout the campaign, and the use of open access tools to disseminate findings. Finalists were also encouraged to build south-south partnerships and seek support from colleagues who were not part of the research team. This finding is consistent with other literature showing that south-south collaboration can improve research quality.^46^

Our study has several limitations. First, the studies identified from the qualitative evidence synthesis were disproportionately from high-income countries and only included articles in the English language. There were however data from some LMICs and the pilot programme did provide detailed complementary information on the feasibility of crowdfunding in LMIC settings. Second, we did not collect information from backers, funders, or other organizations who are important in creating a crowdfunding ecosystem. Third, some researchers do not have a strong background in public engagement which could hinder their ability to conduct robust crowdfunding campaigns. Nevertheless, our pilot shows how researchers with limited social media experience can effectively organize public engagement and successful crowdfunding campaigns.

This research has implications for research and policy (Box 1). Our pilot demonstrates that crowdfunding is feasible to support infectious disease research in LMICs. Public engagement can build horizontal local partnerships, contributing to empowering local funding sources for global health research. From a policy perspective, crowdfunding has not been widely used to support research studies and few platforms focus on scientific and health research. There are also fewer crowdfunding platforms based in LMICs compared to HICs. Global health institutions and universities should help LMIC researchers to consider crowdfunding their research.^2,31^ Our WHO-TDR practical guide provides additional guidance^1^ and helps to expand the uptake of crowdfunding for research. Whilst our initial pilot was organized and supported by TDR Global, further institutional support will be essential for building capacity related to public engagement and crowdfunding.

### Box 1

Practical recommendations for implementing crowdfunding for research *

*1. Public engagement is an important component for conducting a successful crowdfunding for research campaign*

*2. Bi-directional communication may increase the number of crowdfunding donations and enhance the public’s trust and understanding of science*.

*3. Young scholars and early-career researchers may consider crowdfunding for their research*

*4. Smaller crowdfunded grants can top up existing research funding or fund earlystage research that can then be used to apply to public research grants*.

*5. Partnerships with experts can provide some feedback and improve the rigor of research prior to launching a crowdfunding campaign*.

*6. Seeking formal organizational approvals and ethical committee review can increase the likelihood of success*.

*7. To increase donations, campaigns should include quality signals, such as endorsements and testimonials, offer rewards, partner with known NGOs and aim for a realistic funding target*.

*\*A practical guide was developed alongside this review. Researchers interested in strategies and tools to optimise their crowdfunding campaigns can access this guide <u>here</u>*

Our data demonstrate that crowdfunding is an alternative option to support research in LMIC settings and it may be particularly well-suited to early-stage work led by early-career researchers. Crowdfunding has the potential to decentralise research funding and re-orient some of the core underlying principles that underpin global health funding.

## Data Availability

All data produced in the present study are available upon reasonable request to the authors

## Acknowledgments

The authors are grateful to the crowdfunding open call steering committee, judging panel, working group, and end user group. We also wish to thank the internal TDR reviewer, Sassy Molyneux and external peer reviewers, open call participants and finalists. We appreciate support received from Social Entrepreneurship to Spur Health (SESH) and Social Innovation in Health Initiative (SIHI). We would like to thank TDR Global mentors and supporters of this project.

## Contributor statement

BH, JT, and EK developed the original idea for the study. EK, CS, and JT wrote the first draft of the manuscript. EK, CS, PS, SN, and JT were part of the core working group that developed the qualitative evidence synthesis. TW, AW, and MU provided feedback on the evidence synthesis and assessed the practical suggestions. DJ, JA, MH, ML, NJ, PL, WT, and BH were part of the working group that organized the process. All individuals made substantial contributions to the manuscript. All authors reviewed the final version and approved the submission of the manuscript.

