## Supplemental Files for "Crowdfunding for health research: A global systematic review, qualitative evidence synthesis and TDR pilot for LMIC researchers"

Appendix I: In depth details of the open call

Appendix II: Specific topics delivered to finalist at the capacity building workshop

Appendix III Characteristics of included studies

Appendix IV: Updated Search algorithm

Appendix V: Risks in crowdfunding and our mitigation strategies from the TDR pilot

Appendix VI: CASP checklists for all included studies

Appendix VII: PRISMA 2020/ENTREQ Reporting checklist

**Appendix I: In depth details of the open call**

***The crowdfunding open call***

*In partnership with the UNICEF/UNDP/World Bank/WHO Special Programme for Research and Training in Tropical Diseases (TDR), SESH (Social Entrepreneurship to Spur Health) and the Social Innovation in Health (SIHI), the London School of Hygiene and Tropical Medicine) organized the open call for LMIC infectious disease research. The contest was officially launched on May 15^th^, 2019 and the deadline for submissions was June 30^th^, 2019. An open call for submissions was developed and the steering committee reviewed and finalized the details. A simple* [*website*](https://www.seshglobal.org/Crowdfunding) *was created for promotion and to host information on the open call including the purpose, categories of participation, eligibility, timelines, steering committee members, and partners. A submission portal was also available on the website. Participant eligibility included anyone who is a citizen of an LMIC and locally resident in the LMIC where the project is to be implemented.*

***Organizing a steering committee***

*The crowdfunding call had 15 confirmed steering committee members from different countries with, 9 females and 6 males. Of this number, 7 had crowdfunding and/or public engagement research experience in LMICs. The purpose of the steering committee was to provide guidance feedback throughout the duration of the open call. The committee met monthly through 60-minute teleconference meeting. During meetings, issues with regards to the contest design, organization, and implementation and subsequent progress were discussed.*

***Engaging the community to contribute***

*The call for submissions opened on May 16^th^, 2019. Promotional information was disseminated using infographics on social media channels, blog posts, email, and personal contacts. Emails were sent out to partner networks, TDR, SESH, SIHI and to other relevant entities and individuals. Hard copies fliers were printed out with the details on the open call and distributed in relevant places including academic institutions. Promotional activities for entries continued until the deadline for submissions (30^th^ June 2019). At the end of the call, we received a total of 121 unique submissions from 37 different countries.*

***Receiving and evaluating contributions***

*A digital submission form was made using Qualtrics Survey Software and embedded in the contest website. During the call duration, submissions were received via the Qualtrics submission portal on the website and some entries were received through the contest email address. Through the submission form, in addition to the entries, we collected the following sociodemographic details including participants: name, email institution, and country.*

*The evaluation process was conducted in three stages which includes first screening for eligibility, secondly reviewing eligible entries and assigning a score between 1-10. The third stage of evaluation was in-depth review with feedback and comments provided for revisions. At the deadline, all submissions were screened for eligibility using the format for entries posted on the website which includes a 1200-word summary of the project proposed highlighting (1) scientific question and hypothesis; (2) significance of the project; (3) relevance to the public; (4) personal motivation for research and personal connection to the disease and geographic location; (5) areas mentorship will be needed.*

*After screening, the eligible entries were transferred to independent judges for phase 2 judging. A call for volunteer judges to review the submissions was sent out and Our call for volunteer judges received 592 responses and 47 were selected to review submissions.*

*During phase judging, each submission was awarded an individual score between 1 and 10 (where 1 is the weakest and 10 is strongest submission). The scores assigned were based on predetermined criteria set out in a judging rubric. Criteria for evaluation include compelling science, capacity for public engagement, and personal connection to the infectious disease topic. At the end of the second stage judging, five submissions achieved a mean score of 7 and above and emerged as finalists. These 5 finalists in a third stage of judging received detailed feedback on their written pitches.*

***Recognizing finalists***

*The five finalists’ entries were from Guatemala, Sri Lanka, Nigeria, Thailand and Mozambique and included 3 males and 2 females. All the five finalists were announced through the* [*WHO TDR News*](https://www.who.int/tdr/news/2019/tdr-global-challenge-contest-finalists-research-crowdfunding/en/) *and other organizing partner networks and supported to attend a 1.5 day capacity building workshop organized for them in Geneva, Switzerland. During the workshop, the finalists assigned individual mentors and were given time slots to present their project pitches and received group feedback. They also received training on public engagement and storytelling, writing a proposal, crowdsourcing, implementation research, and choosing a platform.*

***Sharing and Implementing solutions***

*During the capacity building workshop, finalists were the given opportunity to share their research projects and receive feedback from the group. After which a working group including some of the SC members and the finalists was set up to meet monthly and further hone the written and video pitches in preparation for campaign launch.*

**Appendix II: Specific topics delivered to finalist at the capacity building workshop**

| **Topics** | **Key messages** |
| --- | --- |
| Crowdfunding and Ethics | Accountability in crowdfunding  Before research: start ethical approval process  During research: giving back to backers, keep updated  After research: open access materials  Other ethical considerations: seek endorsements |
| Storytelling for public Engagement | Capture attention  Be memorable  Inspire action |
| Study design in implementation research | Identifying implementation research problems/questions  Socio-cultural and economic factors  Phase 1: identify possible implementation strategies  Phase 2: Develop implementation strategies  Phase 3: testing effectiveness of implementation strategies  Phase 4: Scaling up |
| Launching a crowdfunding campaign | Expand the network of people engaged with your research.  Communicate your process and results to a broader –and new audience.  Get funding! |
| Project planning | Proposal: scientific merit, social value, well planned project.  Audience should have all the elements to understand the Project to be financed  Different ways to cross the message to the audience.  One way: flow diagram showing the Project development plan  Ideally the diagram should be auto explicit, visual and useful for general audience: |
| Crowdsourcing | Why crowdsourcing to improve health?1) Innovation2) Engages new groups and networks (3) People-centered  Types: challenge contests, hackathons, online collaboration systems  Steps: convene a steering committee, engage the public, receive and judge entries, recognise finalise and the share/implement solutions |
| Writing a Proposal | Illustrating the contribution to impact  Measurable success  Result-based budgeting  Value for money  Risk management |

| Appendix III Characteristics of included studies | | | | | |
| --- | --- | --- | --- | --- | --- |
| SN | **Author name** | **Year** | **Setting** | **Focus** | **Key findings/conclusions** |
| 1 | Sharma et.al., | 2015 | English crowdfunding websites | Simplified content analysis of online crowdfunding campaigns for randomised controlled trial funding | 1. Most crowdfunding campaign funding targets are achieved 2. Crowdfunding is effective to rapidly raise funds for RCTs 3. 95% of campaigns use a flexible model where campaign creators keep all the funds raised 4. Crowdfunding may be useful for pilot or phase 1 studies when funding from government agencies is insufficient. |
| 2 | Aleksina et.al., | 2019 | North America only | Mixed methods analysis of correlates of crowdfunding success in crowdfunding campaigns hosted by Experiment.com (for profit) and Consano (non-profit) **only** | 1. Communication, social networks, and the engagement of a large audience via social media are essential tools for successful crowdfunding 2. Campaigns hosted on Experiment.com were more successful than those on Consano, possibly because Experiment is a larger platform, with a higher number of potential donors. The total funding per project is smaller on Experiment compared to Consano (5000$ more on average). Crowdfunding on platforms with wider publicity are to be preferred, even if they are profit-orientated and charge a fee (Experiment.com) 3. Projects with higher funding targets and those using an innovative approach are less likely to reach their target as donors remain risk averse. 4. Crowdfunding in medical research disregards the disease characteristics and the total value delivered to the society. Researchers can therefore be successful in crowdfunding a wide range of projects. 5. Stated charitable attitudes and behavior cannot be used to forecast actual donation behavior in crowdfunding medical research. |
| 3 | Krittanawong et.al., | 2018 | Top online crowdfunding websites in English (based on site volume) | Simplified content analysis of online crowdfunding campaigns to assess the feasibility of using crowdfunding for cardiovascular research | 1. More than half of crowdfunding campaigns for cardiovascular disease research are unsuccessful. 2. Factors associated with low success included the lack of an easy-to-understand message or campaign video. 3. $5000 to $10 000 is the average amount raised for crowdfunded cardiovascular research. 4. Crowdfunding is most suitable for young investigators looking to conduct pilot studies before applying to larger grants. |
| 4 | Dragojlovic &  Lynd | 2014 | North America and Europe | Simplified content analysis of crowdfunding campaigns for cancer research and rare diseases using mixed methods. | 1. The data suggests that crowdfunding is a viable approach to supporting early-stage proof-of-concept research for both common and rare cancers and for rare inborn genetic diseases. 2. Such an approach could become a valuable additional source of funding for innovators in the drug development arena. Researchers would be well served by splitting their broader research program into a series of smaller discrete projects that could be funded by multiple crowdfunded grants. 3. By crowdfunding early-stage research, innovators might be able to validate their approach, making their projects more competitive in traditional grant competitions and making potential therapeutic interventions more attractive investments for pharmaceutical companies and other investors. 4. Two potential limitations of crowdfunding:  - Rare disease research may be disadvantaged in the crowdfunding arena because donors may have a personal stake in the research may be a driver of donor behaviour - Researchers may not perceive crowdfunding as worth the effort because it requires meticulous planning, time, and long-term attention to building a social media profile.  5. Mitigation strategies – (1) Researchers can partner with experienced foundations who specialise in marketing and fundraising.   (2) University fundraising units are increasingly creating their own crowdfunding portals and will likely be able to provide support and expertise. |
| 5 | Ortiz et.al., | 2018 | North America only | *Amplify Hope* crowdfunding pilot for next generation sequencing funding and mixed methods analysis of the correlates of funding success for rare genetic disease research. | 1. A strong social media network, an active outreach process to networks, as well as engagement within the study all correlated with a higher success rate. 2. Amplify Hope donors were more likely to support projects that were near their fundraising goals, and they found video far more effective for learning about genomics than any other medium. 3. The study hypothesises that the crowdfunding campaign may have increased the public’s knowledge on genetic sequencing and rare diseases. 4. Limitations of crowdfunding included the difficulty of raising funds when the patient population is small, the time and effort dedicated to raising funds, and the need for a strong network and contacts. |
| 6 | Byrnes et.al., | 2014 | North America only | *SciFund* crowdfunding pilot and mixed methods analysis of crowdfunding success factors. | 1. Public engagement and effort on multiple fronts (e-mail, press contact, social media) to engage a large audience are important for funding success. 2. According to qualitative data, scientists doubted that their engagement efforts were successful. 3. Projects usually raise small grants, but there is potential to increase funding amounts. Crowdfunding opens up a new pool of funds for pilot or high-risk projects, allowing a scientist to later leverage their engaged audience alongside preliminary data for larger pools of funds. 4. Many #SciFund projects were on topics that are not normally considered popular with the public – therefore persistent engagement may build an audience for many kinds of projects. 5. #SciFund illustrates that fostering a strong connection between science and society within the culture of academia can benefit both universities and scientists financially and increase public science literacy. 6. To be competitive in the new and dynamic crowdfunding environment, universities must find ways to develop and enrich policies and practices that foster active outreach and engagement by their faculty. |
| 7 | Sauermann et.al., | 2019 | Global (89% of Experiment.com campaigns are US-based) | Standardised content analysis of a sample of Experiment.com projects, with a qualitative analysis of the correlates of funding success | 1. Crowdfunding is used primarily by students and junior investigators, for smaller projects. 2. The success rate of the campaigns in the sample is 48% (higher than NIH grant applications and Kickstarter) 3. Students, junior investigators and women have higher odds of reaching their crowdfunding goals. Results support the view that crowdfunding of scientific research broadens access to resources for groups that have been excluded or disadvantaged in traditional funding systems 4. Projects with higher funding targets have lower odds of success. 5. Projects featuring a video presentation, offering rewards, and those with published lab notes and researcher endorsements have higher odds of success. 6. Conventional signals of quality–including scientists’ prior publications and project risk - have little relationship with funding success, suggesting that the crowd may apply different decision criteria than traditional funding agencies 7. Limitations - the crowd may fund projects that are in legal/political grey zones; crowdfunding side steps traditional peer review; creators may not understand or follow guidelines for ethical research. |
| 8 | Dragojlovic & Lynd | 2016 | North America only | Qualitative survey and analysis of the stated preferences of crowdfunding donors in North America | 1. Respondents indicated a preference for donating to projects conducted by nonprofit research organizations, and an openness to donating to companies that have a ‘for-benefit’ corporate structure. 2. Potential donors were more likely to support non-profit organizations, projects where the university of the lead researcher had an excellent reputation and where other funding was available. Donors also showed a strong preference for projects that have the potential to yield a curative therapy, and that focus on common and paediatric diseases. 3. Donors prioritise treating disease that are deadlier, have more impact on patients’ quality of life, and for which there is a greater unmet need (remember these are stated preferences) 4. Donors prefer donating to friends/family, or with individuals/organisations with whom they already have a relationship. 5. Whilst this study is useful to identify donors’ stated preferences, this may not reflect donors’ actual behavior (see Aleksina et.al.,). |
| 9 | M. Schafer et.al., | 2018 | Global | Standardised content analysis of crowdfunding campaigns on English and German platforms, with a mixed methods analysis of the correlates of crowdfunding success | 1. Presenting strong relevant news factors in project proposals, such as the use of graphical materials (pictures, videos) and humour, is correlated with crowdfunding success 2. One way and especially two-way feedback mechanisms between researchers and backers increase project funding 3. Positive endorsements of the researcher or project and quality signals (academic titles and length of the project description) do not increase funding success. 4. Researcher honors or awards, the promise of rewards and the existence of testimonials are not significant drivers of crowdfunding success. 5. The more information a backer has to relinquish to make a donation, the less successful the project will be. 6. Projects with lower crowdfunding goal are more likely to be successful. 7. Campaigns are more successful on crowdfunding platforms that are focused on scientific projects. No disciplinary field receives less or more in terms of crowdfunding for research. In addition, the amount of views the project gets is not correlated with project success. |

**Appendix IV: Updated Search algorithm**

**Databases:**
PubMed, EMBASE, Web of Science, Scopus, Global Health and Google Scholar.

**Keywords:**

“crowdfunding” OR “crowdfund*

AND

“research”

| **Keyword** | **Search details** | **Results** |
| --- | --- | --- |

| **PubMed** | | |
| --- | --- | --- |
| Crowdfunding | "crowdfunding"[All Fields] | 236 |
| Research | "Research"[MeSH Terms] | 670,259 |
| (crowdfunding) AND (research) | "crowdfunding"[All Fields] AND ("research personnel"[MeSH Terms] OR ("research"[All Fields] AND "personnel"[All Fields]) OR "research personnel"[All Fields] OR "researcher"[All Fields] OR "researchers"[All Fields] OR "research"[MeSH Terms] OR "research"[All Fields] OR "research s"[All Fields] OR "researchable"[All Fields] OR "researche"[All Fields] OR "researched"[All Fields] OR "researcher s"[All Fields] OR "researches"[All Fields] OR "researching"[All Fields] OR "researchs"[All Fields]) | 157 |
| **EMBASE, Web of Science, Scopus, Global Health and Google Scholar** | | |
| Crowdfund* | Crowdfund*.mp. [mp=ti, ab, hw, tc, id, ot, tm, mf, tn, dm, dv, kf, fx, dq, bt, cc, nm, ox, px, rx, an, ui, sy, pt] | 729 |
| Research | Research.mp. [mp=ti, ab, hw, tc, id, ot, tm, mf, tn, dm, dv, kf, fx, dq, bt, cc, nm, ox, px, rx, an, ui, sy, pt] | 15,815,014 |
| Crowdfund*  AND Research | 1 and 2 | 368 |

| *Risks identified* | *Risk mitigation strategies from our TDR pilot* |
| --- | --- |
| *Fraud and deception* | 1) Researchers obtained support from local experts  2) Researchers secured IRB approval prior to crowdfunding launch  3) TDR Global and the researcher’s platform worked in partnership with a crowdfunding platform to facilitate LMIC fundraising |
| *Spreading misinformation* | 1) Researchers employed open access tools to disseminate findings  2) Communicated process and results clearly |
| *Fair allocation of funds* | 1) Researchers included result-based budgeting in the proposal and on the launch page  2) Transparent engagement with backers and the public during the campaign and afterward  3) Illustrating the contribution to impact and measurable success  4) Partnerships with experts |
| *Lack of interest in project* | 1) Researchers created video pitches with storytelling for public engagement which included personal stories  2) Researchers expanded the network of people engaged with their research, built collaborations and mentorship  3) Communicated the process and results to a broader and new audience. |

**Appendix V: Risks in crowdfunding and our mitigation strategies from the TDR pilot**

**Appendix VI: CASP checklists for all included studies**

1. Sharma et al 2015

| A. Validity | 1. Research aim statement | To explore the success of research crowdfunding campaigns by assessing top online (based on site volume) English crowdfunding websites | Y |
| --- | --- | --- | --- |
|  | 2. Qualitative methodology relevant | Yes- Qualitative/descriptive analysis employed with narrative analysis and presentation of finding given here. Descriptive frequencies provided as relevant. | Y |
|  | 3. Appropriate research design | Qual observational article with some basic descriptive frequencies included. Researchers provide a narrative presentation of findings. | Y |
|  | 4. Recruitment strategy appropriate | Data obtained from medical research crowdfunding websites: Experiment, Consano, Petridish, and Cancer Research UK. They searched these crowdfunding websites using the following search terms:” clinical study”,” randomized clinical trial”, and” research”. | Y |
|  | 5. Data collection appropriate | They also independently established whether a campaign met the eligibility criteria of funding for a clinical RCT that was led by an academic or research institution. A consensus process to resolve disagreements was established. | Y |
|  | 6. Relationship between researchers/ participants | One of the researchers had employed crowdfunding methods which could potentially impact on researcher on bias and the influence that researchers could have had on the research. No statement on how bias was addressed in their reporting. | N |
| B. Results | 7. Ethical issues taken into consideration | Ethics not formally applied for or stated in report. Supporting documents showing further data analysis were made available as online appendixes | N |
|  | 8. Data analysis rigorous | Relevant to study topic however evidence is not from primary studies, taking on limitations from original studies | Y/N |
|  | 9. Clear statement of findings | Provides a convincing conclusion, inclusion criteria not clearly stated  Retrieved campaigns from only the top online platforms and limited to English language, rather small number of campaigns selected n=20 | Y |
| C. Usability/ relevance | 10. Value of the research | Correspondence paper, not original research, but valid review results are presented here rather small number of campaigns selected n=20 | Y |

**2. Aleksina et al, 2019** (CASP checklist for qualitative research)

| A. Validity | 1. Research aim statement | Abstract: ‘*To inform researchers applying for this complementary source of research funding, we investigate the determinants of successful crowdfunding campaigns in medical research.’* | Y |
| --- | --- | --- | --- |
|  | 2. Qualitative methodology relevant | Yes – Qualitative methodology is appropriate for this study as the determinants of success are qualitative variables and the data will benefit from qualitative analysis. The research is based on the stated preferences of donors from the Dragojlovic et al study (2016). It shows that none of the attributes found in that 2016 study were in fact useful in predicting the success rate of crowdfunding campaigns. | Y |
|  | 3. Appropriate research design | The decision to use qualitative analysis was not explained. The researcher did however explain why they used ordinary least squares regression to identify success factors.  All the qualitative variables were justified/backed by relevant citations, and a detailed explanation was provided for each one. | Y/N |
|  | 4. Recruitment strategy appropriate | Data comes from the Consano and Experiment.com platforms, which authors explain were chosen after an assessment of the content and suitability of these platforms. The four criteria for inclusion were also clearly provided in the Methods. | Y |
|  | 5. Data collection appropriate | Authors state that ‘data were collected manually’. The methodology of data collection and full data set is made available online. | Y |
|  | 6. Relationship between researchers/ participants | The paper states in the limitations that ‘researchers independently determined the value of the variables in the data set, but that the estimates may be biased. Data was obtained from online sources that were publicly available. Apart from this, there is no reflexion on bias and the influence that researchers could have had on the research. | N |
| B. Results | 7. Ethical issues taken into consideration | No ethics statement is made and there is no attempt to raise issues surrounding consent or confidentiality (however all data was available publicly online) | N |
|  | 8. Data analysis rigorous | The data analysis process is described briefly (OLS regression). There is no theoretical framework but findings are compared to existing evidence. The final sample includes 109 projects from Experiment.com and Consano. Each of the variables included in the data pool are justified and backed up by existing literature or by the Dragojlovic and Lynd et al paper. | Y/N |
|  | 9. Clear statement of findings | The ‘concluding remarks’ at the end of the paper clearly state the research findings, namely the determinants of crowdfunding success. Multiple sources of data were used (triangulation). The findings are discussed in relation to the original research question and are compared to existing evidence.  ‘*We showed that crowdfunding in medical research dis- regards the disease characteristics and the total value delivered to the society. In crowdfunding, scientists with large social networks, either personal or professional, are more likely to achieve their fundraising goal. Scientists who managed to develop good net- working skills and/or their research became covered broadly across the media and have greater chances to succeed. ‘* | Y |
| C. Usability/ relevance | 10. Value of the research | Based in North America only.  The authors discuss how crowdfunding can contribute to current practice (government research funding) and make recommendations on how funding agencies can learn from crowdfunding. There is no discussion on areas for further research or how the findings can be transferred to other populations. | Y |

**3. Krittanawong et al** (CASP checklist for qualitative research) – this was not a pure qualitative research paper, but the same checklist was used for homogeneity purposes

| A. Validity | 1. Research aim statement | ‘our goal was to explore the feasibility of crowdfunding for the support of cardiovascular research’ | Y |
| --- | --- | --- | --- |
|  | 2. Qualitative methodology relevant | A very short article – not qualitative methodology, more descriptive analysis and review of cardiovascular crowdfunded research projects, with a short statistical analysis for correlates of funding success.  In assessing feasibility, a survey among participants may have been a helpful addition. | Y/N |
|  | 3. Appropriate research design | The research design seems appropriate, but in order to assess feasibility, qualitative methods may have been helpful, to get the lived experiences of the researchers completing these CVD crowdfunding campaigns | Y/N |
|  | 4. Recruitment strategy appropriate | Search strategy detailed in the Methods, including search terms and explanation of exclusions/conflicts | Y |
|  | 5. Data collection appropriate | Data collection methods not discussed here – it just states that 3 researchers reviewed the projects included in the study | N |
|  | 6. Relationship between researchers/ participants | Not discussed here. | N |
| B. Results | 7. Ethical issues taken into consideration | No ethics statement/informed consent/confidentiality issues discussed | N |
|  | 8. Data analysis rigorous | Statistical analysis detailed briefly. Lacks a table detailing finding. | Y/N |
|  | 9. Clear statement of findings | Three main findings:   1. Half of crowdfunding campaigns for CVD research are unsuccessful 2. The average amount raised with crowdfunding for CVD research compared with other platforms is $5K to $10K   (Crowdfunding may benefit trainees or young investigators as the average of individuals who are likely to contribute to crowdfunding initiatives is 25 to 34)   3. Crowdfunding may be particularly useful for pilot studies before applying for public research grants, particularly for young investigators | Y |
| C. Usability/ relevance | 10. Value of the research | Findings are compared to available literature and are used to recommend areas for future research. | Y |

**4. Dragojlovic and** Lynd 2014

| A. Validity | 1. Research aim statement | To review the scope and success of existing efforts to crowdfund drug development in oncology and rare diseases, and evaluate the potential for crowdfunding to become a viable source of support for early-stage drug discovery in the future | Y |
| --- | --- | --- | --- |
|  | 2. Qualitative methodology relevant | Yes- Descriptive study and employed narrative synthesis and analysis of finding given here. Descriptive frequencies provided as relevant | Y |
|  | 3. Appropriate research design | Presents descriptive data on 125 crowdfunding campaigns aimed at financing research in oncology (including basic research, drug discovery, and clinical trials). Also describe five campaigns that have succeeded in raising substantial funds | Y |
|  | 4. Recruitment strategy appropriate | They searched all publicly accessible crowdfunding websites to identify all active or expired crowdfunding campaigns related to drug development research listed between 25 October 2013 and 8 November 2013. Exclusion criteria stated in their methods | Y |
|  | 5. Data collection appropriate | Data for this work was retrieved from crowdfunding campaign websites which provides relevant data for the study aim | Y |
|  | 6. Relationship between researchers/ participants | This was not clearly stated on their report. No statement on how this was addressed either | N |
| B. Results | 7. Ethical issues taken into consideration | Yes- the review both on going and past campaigns and excluded campaigns that failed to raise at least 1% of their target from the analysis to avoid ambiguous results | Y |
|  | 8. Data analysis rigorous | Clear presentation of sources of primary data retrieval, high relevance to study topic.  Logical data analysis and presentation given. Sufficient amount of campaigns n=125 across different platforms provides and adequate basis to support the findings and conclusion | Y/N |
|  | 9. Clear statement of findings | Yes- results clearly presented with relevant interpretations of the study findings | Y |
| C. Usability/ relevance | 10. Value of the research | The research findings add to the body of knowledge on the value of crowdfunding a valuable additional source of funding for early-stage researchers and innovators | Y |

**5. Ortiz et. al., 2018** (CASP checklist for qualitative research)

| A. Validity | 1. Research aim statement | Objective is presented in the abstract  5 objectives in the Methods section: *to (1) provide demographic information on the donor population; (2) identify common factors among successful medical crowdfunding campaigns; (3) identify factors that influenced people to donate, as reported by donors; and (4) describe the impact crowdfunding campaigns had on donors’ self-reported knowledge of genomics.*  Needs assessment: *The purpose of all interviews was to establish crowdfunding best practices, elicit recommendations, and develop materials for the training program phase of the study.* | Y |
| --- | --- | --- | --- |
|  | 2. Qualitative methodology relevant | The researchers wanted to get insight from experts and researchers that were successful in conducting their crowdfunding campaign. They used telephone interviews for this which seems relevant. They also used surveys to assess participants/donors’ perspective on the their crowdfunding efforts. Qualitative methodology seems relevant here. | Y |
|  | 3. Appropriate research design | There is no justification on the choice of study design. However the needs assessment was justified and the purpose of the interviews was explained.  The research design seems adequate:  - a needs assessment was conducted through 25 30-minute phone interviews with experts/founders of crowdfunding platforms  - a survey was conducted among participants to assess baseline vs post-crowdfunding knowledge on genomics (only 11 participants overall)  - an anonymous survey was sent to donors after they donated to assess demographics and their knowledge of genomics | N |
|  | 4. Recruitment strategy appropriate | Participant recruitment was detailed. Recruitment documents were made available in supplementary material.  Experts/successful participants were contacted specifically because they had founded crowdfunding platforms or had relevant expertise.  The participants for the crowdfunding trial were recruited through 13,452 emails to rare disease advocacy groups and genetic counsellors, in order to give undiagnosed patients the opportunity to participate. | Y |
|  | 5. Data collection appropriate | The data collection strategy was detailed for the participant and donor surveys (The donor survey was made available in supplementary data). The telephone interview guides were not provided or justified in the main body of text.  Data collection through telephone surveys was appropriate but there seems to have been no standardised method of recording the data. | Y/N |
|  | 6. Relationship between researchers/ participants | No discussion on bias. There is mention of the limitations of using a small patient population (rare diseases) for recruitment. Authors admit that statistical power is limited here. | Y |
| B. Results | 7. Ethical issues taken into consideration | No ethical approval/confidentiality agreement/informed consent explanation. | N |
|  | 8. Data analysis rigorous | The data analysis strategy was not given. No thematic analysis. There is some discussion on potential bias related to the small sample population.  They could have formally assessed the qualitative data collected during the 25 30-minute interviews as these would have provided more insight into what makes a successful campaign from the point of view of experts. | N |
|  | 9. Clear statement of findings | There is clear statement of the findings, including comparison with existing literature on crowdfunding and in relation to the original research question. There was a very limited assessment of credibility (see above)  ‘*We found that social media played an important role in all campaigns. Specifically, a strong social media network, an active outreach process to networks, as well as engagement within the study all correlated with a higher success rate. Amplify Hope donors were more likely to support projects that were near their fundraising goals, and they found video far more effective for learning about genomics than any other medium.’* | Y |
| C. Usability/ relevance | 10. Value of the research | Paper makes recommendation on how this study can contribute to the existing literature on crowdfunding and how it can be used. It also recommends areas for future research. | Y |

**6. Byrnes et. al., 2014** (CASP checklist for qualitative research was used – only the qualitative data was appraised, all quantitative data was excluded if not relevant)

Strong quantitative methods, but lack of formal qualitative analysis of survey results -

| A. Validity | 1. Research aim statement | Based on the #SciFund challenge  3 objectives outlined in introduction: ‘*We therefore set out to ask how the amount of money one could raise via crowdfunding is influenced by:*  *1) building an audience for one's work via science communication*  *2) the amount of effort put into communicating one's science, and*  *3) the different avenues one used to communicate their work*.’ | Y |
| --- | --- | --- | --- |
|  | 2. Qualitative methodology relevant | This study was in fact mixed methods but we focus on the qualitative methodology. The qualitative aspect is justified as the subjective experiences of the SciFund challenge participants is of interest here. The quantitative measures used are also helpful for the study objective. | Y |
|  | 3. Appropriate research design | The authors designed a survey to measure various aspects of crowdfunding from the participants’ perspective. There are specific objectives that are outlined for the survey. | Y |
|  | 4. Recruitment strategy appropriate | The process for recruiting scientists to the challenge was described in detail and there is a table detailing project distribution and the various rounds of the SciFund Challenge. The participants in the final challenge then answered survey questions.  ‘The survey was answered by 47 of the 49 #SciFund round one participants, 48 of 75 round two participants, and 22 of 35 round three participants.’  One outlier project was excuded from the data and this was explained in detail. | Y |
|  | 5. Data collection appropriate | [QUANTITATIVE: data sources are outlined in detail: 1) web visit and donation logs of each crowdfunding project from RocketHub (platform, with dedicated section just for the SciFund challenge) ; 2) publicly available info from the internet (Twitter, Facebook number of likes/tweets)  3) the number of times project videos were viewed (quantitative measures) ]  QUALITATIVE: Survey – the authors provide the full list of questions (based on strategies used to create crowdfunding materials, strategies used to promote campaign, social network size and ongoing online outreach activities) – for all participants of the SciFund challenge. Survey included qualitative questions and quantitative questions.  The survey instrument was updated and this is outlined in the Methods as well: ‘*The survey instrument for rounds two and three differed in some ways from the instrument we used for round one. […]’*  *‘We therefore revised several questions in our survey in order to better assess participant effort for rounds two and three. We were thus able to ask, how does effort modify the effect of audience size on the ability of a researcher to bring people to view their project?’* | Y |
|  | 6. Relationship between researchers/ participants | There Is no discussion on bias here.  Authors declare ‘no competing interests’ – ‘*The organizers of #SciFund were not paid by RocketHub nor did they receive any funds, either directly or indirectly, from #SciFund participants or donors (other than the donor funds Walker, Byrnes, and Faulkes received from their individual projects* (as participants of the challenge)).’  Authors did change the survey across rounds to adapt to the survey responses they received | Y/N |
| B. Results | 7. Ethical issues taken into consideration | No discussion on Ethics/informed consent/confidentiality here | N |
|  | 8. Data analysis rigorous | There is extensive description of the data analysis process for the quantitative analysis - including the hypotheses researchers made and a detailed explanation of the quantitative methods used.  Four stat models (linear models were used) were used to answer four questions:   1) What effect did the number of donors have on crowdfunding success? 2) Where were donations coming from? 3) Was the attention a project received generated from existing social networks or other forms of ‘buzz’ generated by the SciFund campaign itself? 4) Did long term scientific outreach via blogging increase scientists’ outreach-generated social networks   **NO METHODOLOGY ON QUALITATIVE DATA ANALYSIS IS AVAILABLE** and authors only state that the qualitative data was compared to the stat models in order to determine if participant perceptions about crowdfunding success/failure matched the results of stat models. | N |
|  | 9. Clear statement of findings | The quantitative results are well displayed  The qualitative results are briefly explained in the final paragraph, but there is no methodology/framework for data analysis – two Tables (11 and 12) show factors that helped and hurt project fundraising –  The one positive aspect is that qualitative results were compared to statistical analysis quantitative results | N |
| C. Usability/ relevance | 10. Value of the research | Most projects included in analysis were on Conservation biology and ecology (100) (only ~25 were related to health  Extensive discussion on study results and comparing quantitative measures to participants’ perceptions (especially regarding campaign effort)  Some discussion on survey limitations in the Discussion and where the authors fell short  Limited comparison to existing literature  Study explains how this SciFund challenge contributes to current practices in research funding + gives recommendations for the future | Y |

**7.** Sauermann et al 2019

| A. Validity | 1. Research aim statement | To provide new evidence on the state of crowdfunding in scientific research and should be of interest to social scientists as well as to scientists who consider starting their own crowdfunding campaigns. By providing empirical evidence from the specific context of science, this study also contributes to the broader literature on crowdfunding, which tends to focus on general-purpose platforms. | Y |
| --- | --- | --- | --- |
|  | 2. Qualitative methodology relevant | Mixed methods with descriptive analysis. To assess the potential of crowdfunding for scientific research, the authors first reported initial evidence from Experiment.com, They build on this existing work to provide insights into crowdfunding campaigns in an understudied context–scientific research | Y |
|  | 3. Appropriate research design | Mixed methods with descriptive analysis. An initial research was conducted to identify themes ahead of their data collection and analysis | Y |
|  | 4. Recruitment strategy appropriate | They searched all publicly accessible crowdfunding websites to identify all active or expired crowdfunding campaigns related to drug development research listed between 25 October 2013 and 8 November 2013. Exclusion criteria stated in their methods | Y |
|  | 5. Data collection appropriate | Data for this work was retrieved from crowdfunding campaign websites which provides relevant data for the study aim. They provide descriptive information on the creators seeking funding, the projects they are seeking funding for, and features of the crowdfunding campaigns. Then they investigated how these various characteristics are related to campaign success.  They compared the results to prior research on the predictors of fundraising success in crowdfunding but also to research on traditional scientific funding mechanisms such as government grants. And finally examined whether and how predictors of crowdfunding success differ from those that predict attention from a more professional audience–journalists covering scientific research | Y |
|  | 6. Relationship between researchers/ participants | This was not clearly stated on their report. No statement on how this was addressed either | N |
| B. Results | 7. Ethical issues taken into consideration | Yes- permission obtained to share data publicly. IRB application for the work was not reported anywhere in the report. Creator characteristics and project characteristics reported separately. They also provide empirical evidence from the specific context of science, | Y |
|  | 8. Data analysis rigorous | Rigorous data analysis, additional variables were coded based on project description high relevance to study topic. Incomplete data sets were removed, and final campaign data included in analysis was 725 campaigns | Y |
|  | 9. Clear statement of findings | Yes- results were interpreted and a clear statement of their finding presented “Our results highlight significant opportunities for crowdfunding in the context of science while also pointing towards unique challenges. We relate our findings to research on the economics of science and on crowdfunding, and we discuss connections with other emerging mechanisms to involve the public in scientific research” | Y |
| C. Usability/ relevance | 10. Value of the research | The findings highlight important differences between crowdfunding and traditional funding mechanisms for research, including high use by students and other junior investigators but also relatively small project size. This validates some earlier findings published in other crowdfunding studies. this study also contributes to the broader literature on crowdfunding, which tends to focus on general-purpose platforms | Y |

**8. Dragojlovic and Lynd**, 2016 (CASP checklist for qualitative research)

| A. Validity | 1. Research aim statement | Aim: ‘*to help inform the fundraising strategies adopted by biomedical research organizations that are considering the use of crowdfunding, we conducted an online survey of potential North American donors to identify the types of drug development research projects that prospective donors might find most appealing.’* Gap in the literature identified and attempt by the authors to fill it. | Y |
| --- | --- | --- | --- |
|  | 2. Qualitative methodology relevant | Qualitative methodology was appropriate here as the research aimed to collect preferences from individual donors. | Y |
|  | 3. Appropriate research design | Authors did not justify the study design. Presents a very biased picture of donor preferences as they are explicitly asked for these. Stated preferences may deviate from revealed preferences. | N |
|  | 4. Recruitment strategy appropriate | Ipsos USA was in charge of recruitment (a leading survey research firm). A screening questionnaire was used to oversample respondents who had previously donated money to support medical research. Samples included an approximately equal number of male and female respondents. There is limited discussion around recruitment however: we are told that 168 respondents were excluded (with explanation), yielding a usable sample of 814 respondents. | Y |
|  | 5. Data collection appropriate | There is a very detailed explanation of the data collection tool (online survey) and how scoring was done for the responses. There is no justification of the methods used or a discussion on saturation of the data. | Y |
|  | 6. Relationship between researchers/ participants | The authors explain an external company ‘Ipsos’ completed the recruitment of the study. There is limited discussion on potential biases with a reflection on the composition of the Ipsos panel, non-response bias and the exclusion of francophone residents in Canada. | Y |
| B. Results | 7. Ethical issues taken into consideration | No ethical statement or discussion on informed consent/confidentiality | N |
|  | 8. Data analysis rigorous | The data collection strategy and how scores were used was explained in detail in the main body of the research. The data analysis strategy itself is not that detailed. A sample of 814 surveys were used. There is a discussion on bias. | Y |
|  | 9. Clear statement of findings | The findings are explained in detail and are compared to existing literature. | Y |
| C. Usability/ relevance | 10. Value of the research | Directed at North American donors (English-speakers only). There is no discussion on the limitations of the study or on the areas for future research | N |

**9. M. Schafer et al, 2016** (CASP checklist for qualitative research)

| A. Validity | 1. Research aim statement | Abstract ‘*The study at hand identifies and tests explanatory factors influencing the success of scientific crowdfunding projects by drawing on news value theory, the “reputation signalling” approach, and economic theories of online payment.’* | Y |
| --- | --- | --- | --- |
|  | 2. Qualitative methodology relevant | Standardised content analysis, which is a qualitative methodology, was used for this research. The paper uses four theoretical frameworks to organise and analyse the findings (as a conceptual framework to explain crowdfunding success of scientific projects did not exist): ‘news value theory, reputation signalling, theories of online payment and social factors’ | Y |
|  | 3. Appropriate research design | The authors do not discuss the rationale behind the use of standardised content analysis for this research.  However, the research design does seem appropriate – the researchers used specific theoretical frameworks to guide their qualitative analysis. They explain that the codebook they used was organised according to these frameworks. | Y |
|  | 4. Recruitment strategy appropriate | There is a detailed description of the study search, inclusion and exclusion criteria (done by 4 researchers using key terms). There is a table detailing all the crowdfunding platforms included in the analysis and the number of scientific projects per platform. | Y |
|  | 5. Data collection appropriate | There is limited detail on how the data was collected. 371 projects were analysed across 11 crowdfunding platforms. | N |
|  | 6. Relationship between researchers/ participants | No discussion on bias – the authors state that 4 coders participated in coding the projects using specific variables and that intercoder reliability was calculated. | N |
| B. Results | 7. Ethical issues taken into consideration | No ethical statement/confidentiality or informed consent discussion (however all data was available publicly online) | N |
|  | 8. Data analysis rigorous | Standardised content analysis was used. A codebook with 54 variables was created from the data collection. A team of 4 coders coded all the variables for all projects and coding was pretested with intercoder reliability calculated at 0.901 (Holsti). The choice of variables is explained in detail. The ‘Notes’ section at the end provides important insight into data analysis. | Y |
|  | 9. Clear statement of findings | Five models were created to explain the study findings and discuss them in relation to the research aim. The findings are discussed and are compared to existing literature. | Y |
| C. Usability/ relevance | 10. Value of the research | The projects included in this study were mostly on English or German-speaking platforms.  The authors recommend further analysis and recommend further studies on crowdfunding for scientific research, including more explanatory factors and incorporating researcher-specific factors. They also recommend experimental/survey research and recommend modelling the interaction between various factors. | Y |

Appendix VII: PRISMA 2020/ENTREQ Reporting checklist

| Section and topic | Item # | Item guide and description | Location where item is reported |
| --- | --- | --- | --- |
| **Title:**  - Title | 1 | Identify the report as a systematic review | 1 |
| **Introduction** | | | |
| Aim/Rationale | 2 | Describe the aim/rationale for the review in the context of existing knowledge | 4 |
| **Methods** | | | |
| **Synthesis** | 3 | Identify the synthesis methodology or theoretical framework which underpins the synthesis, and describe the rationale for choice of methodology | 4 |
| Eligibility criteria | 5 | Specify the inclusion and exclusion criteria for the review and how studies were grouped for the syntheses. | 5 |
| Information sources | 6 | Specify all databases, registers, websites, organisations, reference lists and other sources searched or consulted to identify studies. Specify the date when each source was last searched or consulted. | 5 |
| Search strategy | 7 | Present the full search strategies for all databases, registers and websites, including any filters and limits used. | 5 & Appendix IV |
| Selection process | 8 | Specify the methods used to decide whether a study met the inclusion criteria of the review, including how many reviewers screened each record and each report retrieved, whether they worked independently, and if applicable, details of automation tools used in the process. | 5 |
| Data collection process | 9 | Specify the methods used to collect data from reports, including how many reviewers collected data from each report, whether they worked independently, any processes for obtaining or confirming data from study investigators, and if applicable, details of automation tools used in the process. | 5 |
| Data items | 10a | Study characteristics- Present the characteristics of the included studies (e.g. year of publication, country, population, number of participants, data collection, methodology, analysis, research questions). | 5 & Table 1 |
|  | 10b | List and define all other variables for which data were sought (e.g. participant and intervention characteristics, funding sources). Describe any assumptions made about any missing or unclear information. | 5 & Table 1 |
| Study risk of bias assessment | 11 | Specify the methods used to assess risk of bias in the included studies, including details of the tool(s) used, how many reviewers assessed each study and whether they worked independently, and if applicable, details of automation tools used in the process. | 5 & appendix VI |
| Effect measures | 12 | Specify for each outcome the effect measure(s) (e.g. risk ratio, mean difference) used in the synthesis or presentation of results. | N/A |
| Synthesis methods | 13a | Describe the processes used to decide which studies were eligible for each synthesis (e.g. tabulating the study intervention characteristics and comparing against the planned groups for each synthesis (item #5)). | 5 |
|  | 13b | Describe any methods required to prepare the data for presentation or synthesis, such as handling of missing summary statistics, or data conversions. | 5 |
|  | 13c | Describe any methods used to tabulate or visually display results of individual studies and syntheses. | 5 & Table 1 |
|  | 13d | Describe any methods used to synthesize results and provide a rationale for the choice(s). If meta-analysis was performed, describe the model(s), method(s) to identify the presence and extent of statistical heterogeneity, and software package(s) used. | 5 |
|  | 13e | Describe any methods used to explore possible causes of heterogeneity among study results (e.g. subgroup analysis, meta-regression). | N/A |
|  | 13f | Describe any sensitivity analyses conducted to assess robustness of the synthesized results. | 5 & Table 2 |
| Reporting bias assessment | 14 | Describe any methods used to assess risk of bias due to missing results in a synthesis (arising from reporting biases). | 5 & appendix VI |
| Certainty assessment | 15 | Describe any methods used to assess certainty (or confidence) in the body of evidence for an outcome. | 5 & Table 2 |
| **Results** | | | |
| Study selection | 16 | Describe the results of the search and selection process, from the number of records identified in the search to the number of studies included in the review, ideally using a flow diagram. | 7 |
| Study characteristics | 17 | Cite each included study and present its characteristics. | 7 & appendix VI |
| Risk of bias in studies | 18 | Present assessments of risk of bias for each included study. | 7 & appendix VI |
| Results of individual studies | 19 | For all outcomes, present, for each study: (a) summary statistics for each group (where appropriate) and (b) an effect estimate and its precision (e.g. confidence/credible interval), ideally using structured tables or plots. | Table 1 and appendix VI |
| Results of syntheses | 20a | For each synthesis, briefly summarise the characteristics and risk of bias among contributing studies. | 8-11 & Table 2 |
|  | 20b | Describe the process for coding of data *(e.g. line by line coding to search for concepts).* | 5 |
|  | 20c | Derivation of themes-Explain whether the process of deriving the themes or constructs was inductive or deductive | 5 & Table 2 |
|  | 20d | Synthesis output- Present rich, compelling and useful results that go beyond a summary of the primary studies (e.g. *new interpretation, models of evidence, conceptual models, analytical framework, development of a new theory or construct).* | 8-11 |
| Reporting biases | 21 | Present assessments of risk of bias due to missing results (arising from reporting biases) for each synthesis assessed. | N/A |
| Certainty of evidence | 22 | Present assessments of certainty (or confidence) in the body of evidence for each outcome assessed. | 8-11 & Table 2 |
| **Discussion** | | | |
| Discussion | 23a | Provide a general interpretation of the results in the context of other evidence. | 12 &13 |
|  | 23b | Discuss any limitations of the evidence included in the review. | 12 |
|  | 23c | Discuss any limitations of the review processes used. | 12&13 |
|  | 23d | Discuss implications of the results for practice, policy, and future research. | 14 |
| **Other information** | | | |
| Registration and protocol | 24a | Provide registration information for the review, including register name and registration number, or state that the review was not registered. | 4 |
|  | 24b | Indicate where the review protocol can be accessed, or state that a protocol was not prepared. | 4 |
|  | 24c | Describe and explain any amendments to information provided at registration or in the protocol. | 4 |
| Support | 25 | Describe sources of financial or non-financial support for the review, and the role of the funders or sponsors in the review. | 15 |
| Competing interests | 26 | Declare any competing interests of review authors. | 15 |
